# Self-reported health 50 years later among Children and Adolescents who experienced The Scorched Earth tactics in Finnmark county, Norway, in World War II, 1944/45

**DOI:** 10.1101/2024.09.19.24313974

**Authors:** Marit Pedersen, Elin Alsaker, Eiliv Lund

## Abstract

**Background:** Most of Finnmark county in Norway was burned down during World War II by the German troops on order from Hitler, the scorched earth tactic. More than 50 000 were evacuated, but 23 000 disobeyed and stayed the winter 1944/45 in caves, turf huts and underneath boats. The purpose of this study was to examine among those being children and adolescents at that time, the effect of place of residence, nutrition and self-reported health consequences of wartime experiences on current self-reported health 50 years later.

**Material and methods:** In connection with the cardio-vascular screening study in Finnmark 1996/97 nine questions regarding health status and experiences during World War II were included in a questionnaire. Altogether 3.471 persons born between 1925 and 1944 responded to the questions: 64% response rate. The dependant variables in the analysis were current self-reported health.

**Results:** In a multivariate analysis those who reported reduction in health due to the war had a higher risk of having poor self-reported health at time of screening; OR=2.7 (95% confidence interval 2.1-3.5). Place of residence 1944/45 in Finnmark was a borderline significant risk factor, while lack of food was associated with reduced self-reported health.

**Conclusion:** Despite the long period of time between World War II and the present screening, a relation between the experience from the war and having a poor self-evaluated health status today was found. The study indicates long-term health consequences of warfare on the civilian population, but due to the cross-sectional design further prospective studies are necessary.

## Background

During World War II the civilian population in almost all European countries were affected. Many were killed, others injured for life (1). According to Statistics Norway 10.262 Norwegians died because of the war, and probably these numbers were higher. It is difficult to find complete numbers because there is no clear line between death during war and reduced length of life due to injuries caused by warfare (2).

In Northern Norway, Finnmark was an area of strategic importance to the German military due to the common border with Russia. Finnmark is the northernmost county in Norway at the same latitude as Alaska, Siberia and Greenland (69-71). During World War II there were more German soldiers in Finnmark than in any other Norwegian county (3). After the Germans failed in the attack towards Russia, the situation for the civilians in Finnmark was dramatically changed to the worse in the autumn of 1944. Adolf Hitler gave order to evacuate the civilian population by force and to burn down all houses (3), a strategy which has been called the scorched earth tactics.

Over 50.000 people in Finnmark and Northern Troms were evacuated by force autumn 1944. The evacuation caused high risk and constrains for many (4). Despite threats of death sentence 23.000 people managed to escape during the evacuation, and they found shelter in caves, turf huts and underneath boats. Being refugees and partisans are two situations known to put people under a lot of pressure during World War II (5). In addition, there were problems concerning food supply and nutrition for those who stayed in Finnmark the winter of 1944/45.

The purpose of this article is to examine what consequences the experiences during the war had on the self-reported health among civilian population in Finnmark 50 years later.

## Material and Methods

The cardio-vascular screening in Finnmark was conducted as a cooperation project between the health service in Finnmark, Norwegian Institute of Public Health and Institute of Community Medicine at the University of Tromsø. The cardio-vascular screening study in Finnmark 1996/97 was the sixth study of this kind in the area, Finnmark VI. In the survey people born in 1925-27 were invited from all communities in Finnmark. People born in 1928-53 were invited from Båtsfjord, Nordkapp, Loppa, Gamvik, Meløy, Kautokeino, Porsanger and Vadsø.

7.836 of the 12.288 invited persons responded to Finnmark VI-survey, a response rate of 64 percent. Of these 3.471 persons born in 1925-1944 were included in this analysis. To compare between the age groups, we chose to divide the respondents into two separate groups, one for those who were adolescents, born 1925-34, or those who were children, born 1935-44, during World War II. We have no information of where in Finnmark the respondents lived during the war.

All participants filled in a three pages questionnaire mailed together with the invitation and brought it with them to the screening. The respondents were asked how they experienced their current health. Answers given were “very good”, “good”, “not so good” and “poor”. In the analysis these four categories were recoded into “very good/good” and “not so good/poor”.

A second questionnaire was distributed at the screening. People either filled it in directly or brought it home together with a prepaid envelope. This questionnaire had nine questions referring to World War II. Place of residence was based on the respondents place of living during the winter of 1944/45. Based on the answers given, the respondents were divided into three groups; 1) Respondents who lived in Finnmark during the war, but were evacuated, 2) Respondents who lived in Finnmark and stayed there over the winter of 1944/45, and 3) Respondents who moved from Finnmark voluntarily or who did not live in Finnmark during the war. Information given in response to where the respondents lived during the war, was adjusted for overlapping answers and not logical combinations. In the statistical analyses people who moved from Finnmark voluntarily or did not live in Finnmark during the war, was used as reference category.

The participants in Finnmark VI were asked whether they had experienced shortage of specific food, shortage of food in general or starvation during the war. These three variables were recoded into one, so common reference category was those who did not experience problems concerning nutrition.

The question about possible health reduction as a consequence of wartime experience were categorised as “yes”, “no” and “do not know”. In the analysis “yes” and “do not know” were recoded into one category, while “no” was used as reference category. Sami ethnicity was defined as two or more grandparents talking Sami language at home. Marital status was grouped as married or not.

## Statistical Analysis

In this analysis we used current self-reported health status as the dependent variable. In the multivariate models the following covariates were used; year of birth, sex, ethnicity, marital status, level of education and economic conditions during childhood. The analysis is conducted using logistic regression with odds ratio and confidence intervals in SPSS 11.0 and estimates with p-values less than 0,05 are considered significant.

## Results

The sample consists of 3.471 persons in Finnmark born between 1925 and 1944 who answered where they were during World War II. Among these 56 percent were born during 1925-34, while 44 percent were born in 1935-44.

Table 1 shows that 1.453 respondents were evacuated from Finnmark and 1.285 respondents stayed in Finnmark during the winter of 1944/45. The 733 respondents who moved from Finnmark voluntarily or did not live in Finnmark during the war was used as reference category. The relationship between year of birth, sex and place of residence were not significant.

**Table 1.** Experiences of war, self-evaluated health due to experiences of war and self-evaluated current health according to birth year and gender.

|  | Born 1925-34 |  | Born 1935-44 |  |  |
| --- | --- | --- | --- | --- | --- |
|  | Males | Females | Males | Females | Total |
| *Place of residences* | N(%) | N(%) | N(%) | N(%) |  |
| *1944/45\** |  |  |  |  |  |
| Was evacuated from Finnmark | 419 (44.0) | 410 (40.6) | 333 (43.3) | 300 (40.1) | 1453 (41.9) |
| Lived in Finnmark winter of 1944/45 | 348 (36.5) | 409 (40.5) | 251 (32.6) | 277 (37.0) | 1285 (37.0) |
| Moved from Finnmark/did not live in Finnmark | 186 (19.5) | 190 (18.8) | 185 (24.1) | 172 (23.0) | 733 (21.1) |
| Total | 7944 (100.0) | 1009 (100.0) | 769 (100.0) | 749 (100.0) | 3471 (100.0) |
| Self-evaluated current health** |  |  |  |  |  |
| Poor | 56 (5.8) | 41 (3.9) | 28 (3.5) | 34 (4.3) | 159 (4.4) |
| Not so good | 418 (43.1) | 517 (48.9) | 286 (35.5) | 322 (41.0) | 1543 (42.6) |
| Good | 452 (46.6) | 432 (40.8) | 424 (52.6) | 369 (46.9) | 1677 (46.3) |
| Very good | 43 (4.4) | 68 (6.4) | 68 (8.4) | 61 (7.8) | 240 (6.6) |
| Total | 969 (100.0) | 1058 (100.0) | 806 (100.0) | 786 (100.0) | 3619 (100.0) |

\* $P < 0.08$ , d.f.=2
\*\* $P < 0.01$ , d.f.=3

Close to each second person responded that the self-evaluated health at the time of the screening was not so good or poor, table 1.

Among those who were either evacuated or stayed in Finnmark during the winter of 1944/45 (table 2), approximately 50 percent reported no reduction in health as a consequence of the war. Of those who moved from Finnmark voluntarily or who did not live in the county during the war, 75 percent reported no effect on the health. More of the population, 12.6%, who stayed in Finnmark during the winter reported that that their health was reduced than among those evacuated, 7.2%, or among those who stayed outside Finnmark, 3.7%.

**Table 2.** Reduction in health due to place of residence 1944/45.

| Place of residence | Reduced health<br>% | Uncertain<br>% | Not reduced health<br>% | Total<br>% | N |
| --- | --- | --- | --- | --- | --- |
| Lived in Finnmark<br>winter of 1944/45 | 12.6 | 40.9 | 47.0 | 100.0 | 1233 |
| Evacuated from<br>Finnmark | 7.2 | 41.8 | 51.0 | 100.0 | 1382 |
| Moved away from<br>Finnmark/did not<br>live in Finnmark | 3.7 | 24.2 | 72.1 | 100.0 | 459 |
| Total * | 8.8 | 52.6 | 38.6 | 100.0 | 3074 |

\* $P < 0.01$ , d.f.=4

Table 3 shows the relationship between the place of residence during 1944/45 and current self-reported health. Significant more people staying in Finnmark had poor or not so good health.

**Table 3.** Reduction in current self-reported health due to place of residence.

| Place of residence | Poor/not good<br>% | Good/very good<br>% | % | N |
| --- | --- | --- | --- | --- |
| Lived in Finnmark<br>winter of 1944/45 | 50.0 | 50.0 | 100.0 | 1284 |
| Evacuated from<br>Finnmark | 48.2 | 51.8 | 100.0 | 1452 |
| Moved away from<br>Finnmark/did not<br>live in Finnmark | 41.3 | 59.7 | 100.0 | 732 |
| Total * | 47.2 | 52.8 | 100.0 | 3468 |

\* $P < 0.01$ , d.f.=2

Reduced health as a consequence of war time experiences was a significant risk factor for reduced self-reported health at time of screening; OR 2.7 (95% CI: 2.1-3.5) (table 4). In the mutually adjusted analysis place of residence was only borderline significant. Shortage of food was a significant predictor, while starvation showed no effect on self-reported health.

**Table 4.** Odds ratio (OR) with 95% confidence interval (95% CI) for not so good/poor self-evaluated current health.

| Variable | OR* | 95% CI |
| --- | --- | --- |
| **Health conditions due to war** |  |  |
| No reduced health | 1.0 | (Reference) |
| Reduced or uncertain of reduced health | 2.7 | 2.1-3.5 |
| **Place of residence** |  |  |
| Moved from Finnmark or did not live in Finnmark | 1.0 | (Reference) |
| Lived in Finnmark and was evacuated | 1.3 | 0.9-2.0 |
| Stayed in Finnmark winter of 1944/45 | 1.4 | 0.9-2.2 |
| **Nutrition during the war** |  |  |
| No problems concerning nutrition | 1.0 | (Reference) |
| Shortage of specific kind of food | 1.2 | 0.8-1.8 |
| Shortage of food during the war | 1.7 | 1.1-2.6 |
| Starved | 1.2 | 0.6-2.2 |

\* Mutually adjusted for the other factors in the table, and in addition gender, age, marriage, year of education and living conditions in the childhood and talking about of experiences of war.

## Discussion

Our findings give reason to believe that war experiences in childhood and adolescent could have affected this specific generation’s later health condition. Respondents who was evacuated or lived in Finnmark during the winter of 1944/45 experienced reduction in health status as a consequence of the war, a reduction which showed a relationship to current self-reported health.

### Weakness and strength

We believe that the questions concerning place of living were more easily to recall than the questions concerning nutrition. Nevertheless, we believe that the experiences of war affected people so much that most of them remember them well. The number of evacuated and those who stayed in Finnmark the winter of 1944/45 in this study reflect the actual number of people who experienced this (3, 4, 6). This indicates sufficient extern validity, and we believe this material is representative for the population in Finnmark. In the analysis we adjusted for known risk factors for self-reported health in the regression analyses (7,8,9,10).

A major problem with this cross-sectional analysis is the possibility of reversed causality. People who have a reduced self-reported health could place this on their previous wartime experiences. The participants in this study first answered the question about self-reported health, and the questions on wartime experiences was answered in the second questionnaire. The hypothesis of the study was not told to the participants, and the focus in the public on this hypothesis was at that time small.

### The Evacuation

The Germans believed that 40 percent of the civilian population in Finnmark would not survive the evacuation, the Norwegian nazi’s believed 25 percent would die. Both of these calculations exceeded the number of deaths. The good weather during the autumn 1944 may be the reason for this (6). Still there were people dying during the evacuation of Finnmark and Northern Troms. At the time of the evacuation about 100 people had to be buried in other places. Most of the population in Nesseby, Sør- and Nord-Varanger avoided evacuation since the German evacuation-authorities had to escape from Finnmark to get away from the Russian army.

Those who stayed in Finnmark and those who were evacuated, more often reported that their health was reduced by the war. We do not have information concerning what these health problems are, but from other studies we know that circumstances during the evacuation could be terrible (3, 6). “Karl Arp”, a German carrier, sailed from Indre Billefjord November 10^th^ 1944. Onboard there were 1.900 evacuated from Tana, Vadsø and Porsanger. The trip lasted for five days, and before they reached Narvik 95 percent had dysentery, typhus and other infection diseases. The evacuated were often left without having any communication with relatives and friends who had escaped the evacuation. Some claim that the most pathogenetic factor during the war was anxiety, and especially the fear for what happened to family and friends (11).

### The Caves

November 30^th^ 1944, head of defence, Crown Prince Olav, talked through radio from exile in London. The head of defence said the population in Finnmark should not obey the enemies order of evacuation, but instead they should hide in secure places when the evacuation was about to take place. Approximately 23.000 people did what the head of defence asked them to do. Staying in Finnmark winter of 1944/45 lead to solidarity, but also to anxiety, diseases and shortage of food (4).

Living in caves was traumatic for lot of people. On Sørøya in West-Finnmark the conditions became terrible during the winter and several thousands of people were evacuated under dramatic circumstances by the English fleet. About 4.000 people hide in a mine in Bjørnevatn in Sør-Varanger the autumn of 1944. The food consisted of salt fish, replacement for flour, coffee replacement and whale blubber as replacement for butter. Potatoes were not accessible.

### Nutrition during World War II

The changes in incidence and mortality from various diseases during the World War II in Norway were considerable (12, 15). Health is dependent on qualitative and quantitative sufficient nutrition. The number of calories was only barely reduced in Norway during World War II (2). In 1942 daily number of calories in rationalized goods except vegetables and fish, was down in 1.501 calories. An adult needs approximately 2.500 calories daily. As the time of the occupation became longer, more goods were being rationalized, at the same time as rations became smaller and the quality of them poorer. However, small children, sick people and pregnant women were allowed larger rations then others (2). Only 172 respondents, or 5 percent, answered that they starved during the war, but many experienced shortages of food or shortage of certain kind of food. In this study we found that among those who were 1-9 years old in 1944, 23 percent did not experience starvation, insufficient amount of food or shortage of specific kind of food during the war, compared to 9 percent of those who were 10-19 years old. Nutrition problems in the oldest age group could have effect in this cohort’s ability to grow (12,15,16,17).

### Self-evaluated current health

Those who had health problems due to the war had almost three times higher risk than others for experiencing their current health as reduced. This relationship is adjusted for age, sex, ethnicity, conditions during childhood and socio-economic factors. Those who experienced nutritional problems during the war also reported more often reduced health. Those who experienced war, and were born in 1924-44, was at the time of the survey between 52 and 71 years old.

In this study we do not have information regarding what kind of health problems the participants experience due to the war. But it is likely that the different constrains; evacuation or staying in Finnmark winter of 1944/45 led to different challenges to peoples health. The long interval between the wartime experiences and the screening is an analytical problem. If the hypothesis of reduced health determined by wartime experiences should be correct, then the lack of a prospective design could lead to a “healthy survivor” effect. In our study there are more females than males who evaluated their own health as poor. In the study “Kvinner og kreft” women living in Northern Norway in 1991/92 had higher prevalence of poor self-evaluated health compared to women elsewhere in Norway (1), but these discrepancies disappeared after adjustment for socio-economic factors.

### Long-term health consequences

Several hypothesis link poor nutrition and living conditions in utero, early childhood or puberty to later increase in risk both for cardiovascular diseases (16,17) and cancer (19). This analysis indicates that persons who were in utero or early childhood only to a smaller extent reported starvation or lack of food in contrast to those in puberty or young adolescents.

## Conclusion

These findings from a cross-sectional analysis should be verified in a prospective design.

## Data Availability

Due to legal ethical restrictions on the dataset, which contains potentially sensitive information, the data could be made available upon request to the National Institute of Public health, Oslo, Norway.

## Abbreviations not used

## Declarations

### Ethical approval and consent to participate

The cardio-vascular screening study in Finnmark 1996/97 was run by the National Institute of Health. The institute was responsible for ethical clearance. All participants gave written informed consent. The information used in this analysis was group anonymous and as such do not need ethical clearance for this table analyses according to Norwegian Law of Health research paragraph 20.

### Consent for publication

Not applicable.

### Availability of data and materials

Due to ethical restrictions on the dataset, which contains potentially sensitive information, the data could be made available upon request to the National Institute of Public health, Oslo, Norway.

### Authors’ contribution

MP did the statistical analyses and wrote the manuscript. EA participated in data handling and statistical analyses. EL initiated the study and conducted study supervision. All authors contributed toward data analysis, drafting and critically revising the manuscript. All gave final approval of the version to be published. MP and EA are presently not working at UiT The Arctic University of Norway.

## Acknowledgement

We are thankful for the cooperation with the National Institute of Health. We are thankful and impressed by the participation from the local populations.

## Disclaimer

The national Institute of Health is not responsible for the analysis or interpretations of the data presented.

## References

1. Eitinger L, Vold O, Weisæth L. Krigsskader og senvirkninger. Krigspensjonering gjennom 50 år. [Injuries sustained in war and long-term consequences. War pensions through 50 years]. Oslo: Rikstrygdeverket 1995.

2. Hjeltnes G. Norge 1940-45. Hverdagsliv i krig. [Day to day life during war]. Oslo: Aschehoug 1987.

3. Finstad BP. Tvangsevakueringa av Finnmark og Nord-Troms høsten 1944. [The forced evacuation of Finnmark and Nord-Troms autumn 1944]. In Mellem R. Så jaga dem oss fra heiman våres. Stamsund: Orkana forlag 1998.

4. Gjenreisningsmuseet: http://museumsnett.no/gjenreisningsmuseet/innledning.htm [The Museum of Reconstruction of Finnmark and Nord-Troms]. (10.9.2003).

5. Norges Offentlige Utredninger. Alta bataljon. [Official reports to the Government. Alta battalion].NOU 1998:12. Oslo: Statens forvaltningstjeneste/Statens trykning 1998.

6. Westrheim H. Landet de brente. Tvangsevakueringen av Finnmark og Nord-Troms høsten 1944. [The land they burnt. Evacuation of Finnmark and Nord-Troms autumn 1944]. Oslo: Nordnorsk forfatterlag/Tiden Norsk forlag 1978.

7. Skretting Lunde E. De fleste eldre vurderer helsa positivt. [Most elderly evaluate their health as good]. Samfunnspeilet 2000: 35–40.

8. Krüger Ø. Helse i Norge – hva kan befolkningsundersøkelser fortelle oss? .[Health in Norway - what can population-studies tell us?] in Larsen Ø, Alvik A, Hagestad K, Nylenna M. Helse for de mange – Samfunnsmedisin i Norge. Oslo: Gyldendal Norsk forlag 2003.

9. Lund E, Kumle M, Sandaune AG. Flyttemønsterets betydning for selvvurdert helse blant norske kvinner i alderen 24-69 år. [Significance if migration patterns in self-reported health among Norwegian women aged 34-69 years]. Tidskr Nor Lægeforen 1998; 24: 3752–5.

10. Lund E. Sosioøkonomisk status, selvvurdert helse og sykdom blant norske kvinner i alderen 45-64 år. [Socio-economic status, self-reported health and morbidity among Norwegian women aged 45-64]. Tidskr Nor Lægeforen 2000; 10: 1131–5.

11. Lønnum A. Helsesvikt – en senfølge av krig og katastrofe. [Health failure – a long-term consequences of war and disaster]. Oslo: Gyldendal Norsk forlag 1969

12. Strøm A. The influence of wartime conditions in Norway. Oslo: Akademisk Trykningssentral 1954.

13. Strøm A. Krig og helse. [War and Health]. Oslo: H. Aschehoug & Co. 1974

14. Forsdahl I. Helseforholdene i Sør-Varanger i krigsårene 1940-44. [Health conditions in Sør-Varanger 1940-44]. Tidskr Nor Lægeforen 1990; 17: 2192–7.

15. Forsdahl A. Sundhetstilstanden, hygieniske og sosiale forhold i Sør-Varanger 1869-1975 belyst ved medisinalberetningene. [Health, hygenic and social conditions in Sør-Varanger 1869-1975 studied by the medical-reports]. Tromsø: Institutt for samfunnsmedisin 1976.

16. Godfrey KM, Barker DJP. Fetal nutrition and adult disease. Am J Clin Nutr 2007; 71 (suppl): 1344S–52S.

17. Forsdahl A. Are poor living conditions in childhood and adolescence an important risk factor for arteriosclerotic heart disease? Br J Prev Soc Med 1977; 31; 91–5.

18. Svensson E, Grotmol T, Hoff G, Langmark F, Norstein J, Tretli S. Trends in colorectal cancer incidence in Norway by gender and anatomic site: an age-period-cohort analysis. Eur J Cancer Prev 2002; 11; 489–95.

